# Outcomes of SARS-CoV-2 Infection in Patients with Chronic Liver Disease and Cirrhosis: a N3C Study

**DOI:** 10.1101/2021.06.03.21258312

**Authors:** Jin Ge, Mark J. Pletcher, Jennifer C. Lai, for the N3C Consortium

## Abstract

**Background and Aims:** In patients with chronic liver diseases (CLD) with or without cirrhosis, existing data on the risk of adverse outcomes with SARS-CoV-2 infection have been mixed or have limited generalizability. We used the National COVID Cohort Collaborative (N3C) Data Enclave, a harmonized electronic health record (EHR) dataset of 5.9 million nationally-representative, diverse, and gender-balanced patients, to describe outcomes in patients with CLD and cirrhosis with SARS-CoV-2.

**Methods:** We identified all chronic liver diseases patients with and without cirrhosis who had SARS-CoV-2 testing documented in the N3C Data Enclave as of data release date 5/15/2021. The primary outcome was 30-day all-cause mortality. Survival analysis methods were used to estimate cumulative incidences of death, hospitalization, and mechanical ventilation, and to calculate the associations of SARS-CoV-2 infection, presence of cirrhosis, and demographic and clinical factors to 30-day mortality.

**Results:** We isolated 217,143 patients with CLD: 129,097 (59%) without cirrhosis and SARS-CoV-2 negative, 25,844 (12%) without cirrhosis and SARS-CoV-2 positive, 54,065 (25%) with cirrhosis and SARS-CoV-2 negative, and 8,137 (4%) with cirrhosis and SARS-CoV-2 positive. Among CLD patients without cirrhosis, 30-day all-cause mortality rates were 0.4% in SARS-CoV-2 negative patients and 1.8% in positive patients. Among CLD patients with cirrhosis, 30-day all-cause mortality rates were 4.0% in SARS-CoV-2 negative patients and 9.7% in positive patients.

Compared to those who tested SARS-CoV-2 negative, SARS-CoV-2 positivity was associated with more than two-fold (aHR 2.43, 95% CI 2.23-2.64) hazard of death at 30 days among patients with cirrhosis. Compared to patients without cirrhosis, the presence of cirrhosis was associated with a three-fold (aHR 3.39, 95% CI 2.96-3.89) hazard of death at 30 days among patients who tested SARS-CoV-2 positive. Age (aHR 1.03 per year, 95% CI 1.03-1.04) was associated with death at 30 days among patients with cirrhosis who were SARS-CoV-2 positive.

**Conclusions:** In this study of nearly 220,000 CLD patients, we found SARS-CoV-2 infection in patients with cirrhosis was associated with 2.43-times mortality hazard, and the presence of cirrhosis among CLD patients infected with SARS-CoV-2 were associated with 3.39-times mortality hazard. Compared to previous studies, our use of a nationally-representative, diverse, and gender-balanced dataset enables wide generalizability of these findings.

## Introduction

Hepatic involvement is common in Severe Acute Respiratory Syndrome Coronavirus 2 (SARS-CoV-2) infection and the COVID-19 disease with clinical manifestations ranging from asymptomatic liver function test elevations to acute hepatic decompensations.^1–4^ In patients with existing chronic liver diseases (CLD) and cirrhosis, however, the outcomes of SARS-CoV-2 infection have been mixed.^5–10^ Previous small-scale studies from tertiary referral centers have demonstrated mortality rates approaching 40% for patients with cirrhosis who were infected by SARS-CoV-2.^7,10^ Other studies, however, have shown that patients with cirrhosis who test positive for SARS-CoV-2 infection had similar mortality rates compared to those patients hospitalized with complications of cirrhosis without SARS-CoV-2 infection.^9^

A recent study of patients with and without cirrhosis based on national data extracted from the United States Department of Veterans Affairs Clinical Data Warehouse demonstrated that patients with cirrhosis were less likely to test positive for SARS-CoV-2 but when positive were approximately 3.5 times more likely to die from all-causes compared to those who tested negative. While this was one of the largest studies of outcomes of SARS-CoV-2 infection in patients with cirrhosis to date, the underlying patient population was 88% male, limiting generalization to other patient populations.^11^

The National COVID Cohort Collaborative (N3C) was formed in April 2020 as a centralized resource of harmonized electronic health record (EHR) data from multiple health systems around the United States to accelerate understanding of SARS-CoV-2 infection.^12,13^ As of May 15, 2021, 208 clinical sites had signed data transfer agreements and 56 sites had harmonized data included in the N3C Data Enclave. The N3C Data Enclave provides a diverse and nationally representative central repository of harmonized EHR data and represents a new model for collaborative data sharing and analytics. The initial results up to December 2020 from the N3C main cohort has been previously characterized and described.^14^

To address the conflicting results and various gaps of previous studies, we used data from N3C Data Enclave to answer three distinct questions regarding outcomes of SARS-CoV-2 infection in CLD patients:

1. What is the association between SARS-CoV-2 and mortality in CLD patients with cirrhosis?
2. What is the association between cirrhosis and mortality in CLD patients who tested positive for SARS-CoV-2?
3. What are the factors associated with mortality among CLD patients with cirrhosis who tested positive for SARS-CoV-2?

## Methods

### The National COVID Cohort Collaborative (N3C)

The N3C is a centralized, curated, harmonized, secure, and nationally representative clinical data resource with embedded analytical capabilities. The N3C is comprised of members from the National Institutes of Health (NIH) Clinical and Translational Science Awards (CTSA) Program and its Center for Data to Health (CD2H), the IDeA Centers for Translational Research, the National Patient-Centered Clinical Research Network, the Observational Health Data Sciences and Informatics network (OHDSI), TriNetX, and the Accrual to Clinical Trials network. N3C’s design, infrastructure, deployment, and initial analyses from the main N3C cohort have been previously described. The analytics platform “N3C Data Enclave” is a secure cloud-based implementation of Palantir Foundry (Palantir Technologies, Denver, Colorado) analytic suite hosted by the NIH National Center for Advancing Translational Sciences (NCATS).^12,14^

The full N3C Data Enclave includes EHR data of patients from partner sites who were tested for SARS-CoV-2 or had related symptoms after January 1, 2020. For patients included in the N3C Data Enclave, encounters in the same source health system beginning on or after January 1, 2018 are also included to provide lookback data. N3C utilizes centrally maintained “shared logic sets” for common diagnostic and phenotype definitions.^12,14^ All EHR data in the N3C Data Enclave are harmonized in the Observational Medical Outcomes Partnership (OMOP) common data model, version 5.3.1.^15,16^ In the OMOP common data model, classification vocabularies, such as International Classification of Diseases, Tenth Revision, Clinical Modification (ICD-10-CM), or Standard Nomenclature of Medicine (SNOMED); are incorporated and mapped to standard OMOP concepts based on semantic and clinical relationships.^17^ Vocabulary classification and mapping of various ontologies to the OMOP standard vocabulary is maintained by OHDSI and publicly available on ATHENA (http://athena.ohdsi.org/), which is a web-based vocabulary repository.^18^ For all analyses, we utilized the de-identified version of the N3C Data Enclave, versioned as of May 15, 2021 and accessed on May 20, 2021. To protect patient privacy, all dates in the N3C Data Enclave are uniformly shifted up to plus/minus 180 days within each partner site in the de-identified database.

### Definition of SARS-CoV-2 Status

SARS-CoV-2 testing status was based on the N3C shared logic set.^12,14^ Specifically, OMOP concept identifiers signifying antibody, culture, or nucleic acid amplification testing for SARS-CoV-2 (Supplemental Table 1) were queried among all patients included in the N3C Enclave. The “index date” for all analyses was defined as the date of the earliest positive test (for SARS-CoV-2 positive patients) or earliest negative test (for SARS-CoV-2 negative patients).^11^ Patients who underwent repetitive SARS-CoV-2 testing were, therefore, classified in either positive or negative categories based on the above definitions governing the earliest test. Patients who did not have SARS-CoV-2 testing by the above definitions (e.g., those who were diagnosed with “suspected COVID-19” and did not undergo testing) were excluded from all analyses. To account for uniform date-shifting that occurs per partner site in the de-identified N3C Data Enclave, we calculated a “maximum data date” to reflect the latest known date of records for each data partner site and excluded all patients who were tested within 90 days of the “maximum data date” at each partner site.

### Definitions of Chronic Liver Disease and Cirrhosis

CLD diagnoses were made based on documentation of at least one standard OMOP concept identifier corresponding to previously validated ICD-10-CM codes for liver diseases (Supplemental Table 2) at any time prior to the index date.^19–22^ As “steatosis of the liver” is a common finding in either alcohol-associated liver disease (AALD) and non-alcoholic fatty liver disease (NAFLD), patients with OMOP concept identifier 4059290 (corresponding to ICD-10-CM code K76.0) and at least one OMOP concept identifier describing alcohol use or dependence (Supplemental Table 2) were categorized as those with AALD in accordance with definitions by the Centers for Disease Control and Prevention (CDC) and the National Institute on Alcohol Abuse and Alcoholism Alcohol Epidemiologic Data System.^23–26^ Patients with OMOP concept identifier 4059290 without an alcohol use OMOP concept identifier were categorized as NAFLD.

Diagnoses were determined in a hierarchical manner such that NAFLD categorization was made only after exclusion of all other causes of CLD. In those patients identified to have CLD, diagnoses of cirrhosis were made based on documentation of at least one OMOP concept identifier corresponding to previously validated ICD-10-CM codes for cirrhosis and its complications (Supplemental Table 2) at any time prior to the index date.^11,27^ Diagnoses of cirrhosis, therefore, can only take place in the setting of an existing CLD diagnosis. Patients who had undergone orthotopic liver transplantation as signified by OMOP concept identifier 42537742 (corresponding to ICD-10-CM code Z94.4) were excluded from all analyses.

### Study Design and Questions of Interest

Using the above definitions for SARS-CoV-2 testing and chronic liver disease/cirrhosis; we isolated our adult patients (with age >= 18 years documented) study population. We divided the study patients into four cohorts (Figure 1):

**Figure 1.**
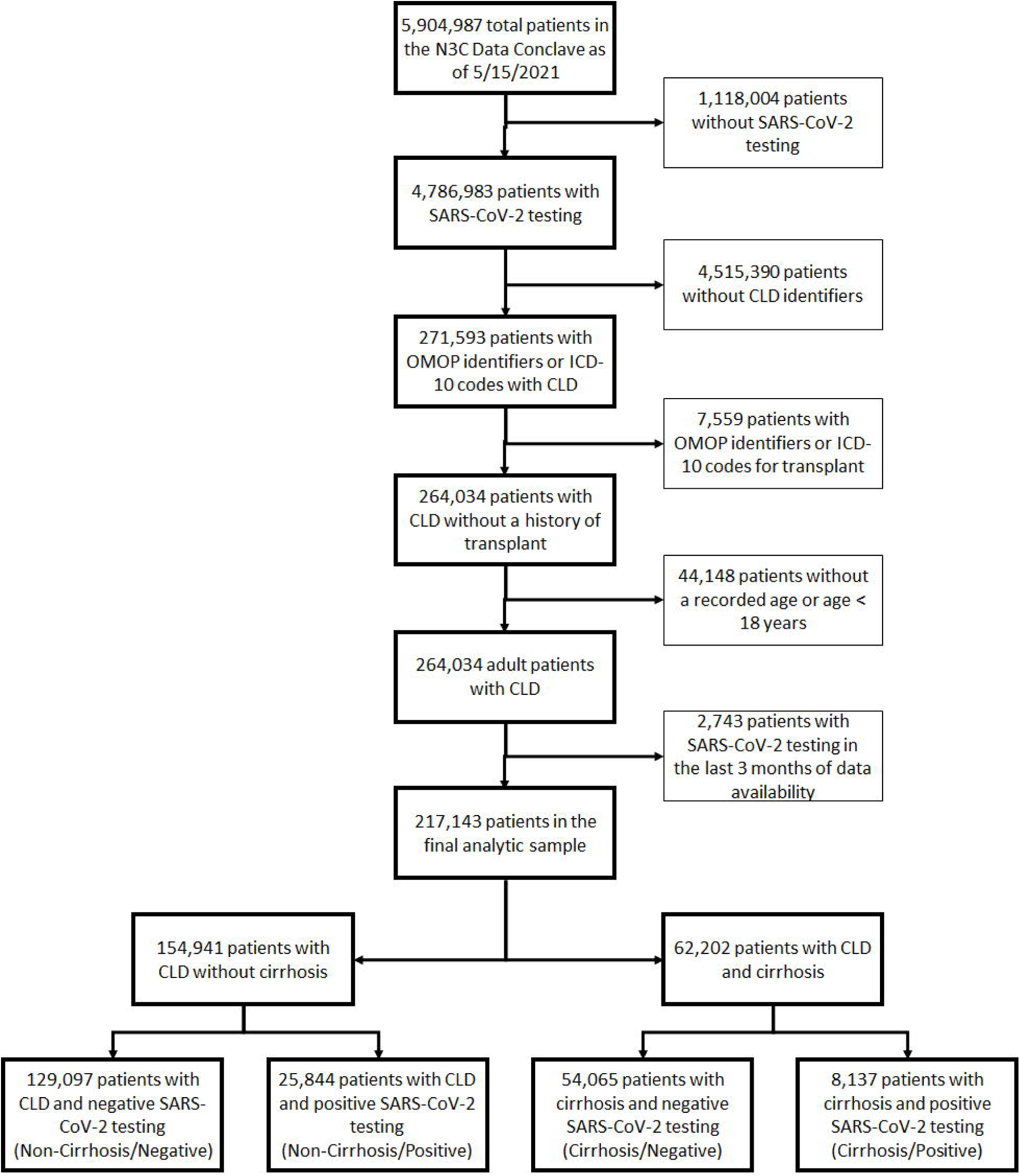
Isolation of CLD patients with and without cirrhosis from the main N3C cohort.

- CLD without cirrhosis and SARS-CoV-2 negative – “Non-Cirrhosis/Negative.”
- CLD without cirrhosis and SARS-CoV-2 positive – “Non-Cirrhosis/Positive.”
- CLD with cirrhosis and SARS-CoV-2 negative – “Cirrhosis/Negative.”
- CLD with cirrhosis and SARS-CoV-2 positive – “Cirrhosis/Positive.”

Based these cohorts, we investigated three questions or associations of interest concerning SARS-CoV-2 infection in CLD patients with or without cirrhosis:

1. What is the association between SARS-CoV-2 and all-cause mortality at 30 days in CLD patients with cirrhosis? This a comparison between CLD patients with cirrhosis who tested positive for SARS-CoV-2 (“Cirrhosis/Positive”) and CLD patients with cirrhosis who tested negative for SARS-CoV-2 (“Cirrhosis/Negative”).
2. What is the association cirrhosis and all-cause mortality at 30 days in CLD patients who tested positive for SARS-CoV-2? This is a comparison between CLD patients with cirrhosis who tested positive for SARS-CoV-2 (“Cirrhosis/Positive”) and CLD patients without cirrhosis who tested positive for SARS-CoV-2 (“Non-Cirrhosis/Positive”).
3. What are the demographic and clinical factors associated with all-cause mortality at 30 days among CLD patients with cirrhosis who tested positive for SARS-CoV-2 (“Cirrhosis/Positive”).

### Outcomes

All patients were followed until their last recorded visit occurrence, procedure, measurement, observation, or condition occurrence as per the OMOP data model in the N3C Data Enclave. The primary outcome was all-cause mortality at 30-days after the index SARS-CoV-2 test date. Secondary outcomes investigated included hospitalization on the index date or within 30- and 90-days after the index date, mechanical ventilation on the index date or within 30- and 90-days, and all-cause mortality at 90-days after the index date. The outcome of death was ascertained based on EHR data indicating in-hospital death, out-of-hospital death, or referral to hospice at each site. The outcome of mechanical ventilation was ascertained by OMOP procedure or condition concepts based on standardized terminology code. The outcome of hospitalization was ascertained based on recorded OMOP visits concepts. These outcomes were defined centrally and queried based on concept sets defined in N3C shared logic and implemented on the full N3C cohort.^12,14^ To account for potential delays in data reporting/harmonization and outcome ascertainment from data partner sites, we had excluded all patients who had SARS-CoV-2 testing within 90 days of the “maximum data date” as defined above.

### Baseline Characteristics

Baseline demographic characteristics extracted from N3C Data Enclave included age, sex, race/ethnicity, height, weight, body mass index, and state of origin. States of origin were classified into four geographic regions (Northeast, Midwest, South, and West) defined by the CDC’s National Respiratory and Enteric Virus Surveillance System (NREVSS).^28^ Patients were categorized as living in “Other/Unknown” region of origin if they originated from territories not otherwise classified (e.g. Guam, Puerto Rico, United States Virgin Islands, and etcetera) or if their state of origin was not known due to censoring to protect patient privacy in zip codes with few residents. Comorbid conditions were extracted based on N3C shared logic sets for calculation of Charlson Comorbidity Index (CCI) score to estimate each patient’s overall burden of comorbidity in the full N3C cohort.^14^

Components of common laboratory tests (basic metabolic panel, complete blood count, liver function tests, and serum albumin) were extracted based on N3C shared logic sets except for international normalized ratio (INR), which we custom-defined based on standard OMOP concept identifiers encompassing this measurement (Supplemental Table 3). We extracted the most complete values to calculate the model for end-stage liver disease-sodium (MELD-Na) score closest to or on the index date from 30 days before the index date or within 7 days after the index date. 56% of patients had laboratory tests were performed within 2 days of the index date available. 17,460 patients, which represented 8% of the full analytical sample, had full laboratory data for calculation of MELD-Na scores. The time frame of 30 days before or 7 days after the index date was consistent with definitions used by N3C to identify hospitalizations of interest in the main cohort characterization.^14^

### Statistical Analyses

Clinical characteristics and laboratory data were summarized by medians and interquartile ranges (IQR) for continuous variables or numbers and percentages (%) for categorical variables. Comparisons between groups were performed using chi-square and Kruskal-Wallis tests where appropriate. We used the Kaplan-Meier method to calculate 30-day and 90-day cumulative incidences of complications of SARS-CoV-2 infection among each cohort: hospitalization, mechanical ventilation, and death. We used Cox proportional hazard models to evaluate the associations between SARS-CoV-2 and mortality among patients with cirrhosis, between cirrhosis and mortality among CLD patients who tested positive for SARS-CoV-2, and factors with mortality for Cirrhosis/Positive patients. In all multivariable analyses, we adjusted for age, sex, race/ethnicity, CLD etiology, CCI score, and region of origin.

As full MELD-Na scores and serum albumin values were available for 17,460 (8%) and 73,313 (34%) patients in the analytical sample, we separately conducted sensitivity analyses of the above models comparing patients with cirrhosis with additional adjustments. Two-sided p-values < 0.05 were considered statistically significant in all analyses. Data queries, extractions, and transformations of OMOP data elements and concepts in the N3C Data Enclave were conducted using the Palantir Foundry implementations of Spark-Python, version 3.6, and Spark-SQL, version 3.0. Statistical analyses were performed using the Palantir Foundry implementation of Spark-R, version 3.5.1 “Feather Spray” (R Core Team, Vienna, Austria).^29^

### Institutional Review Board Oversight

Submission of data from individual centers to N3C are governed by a central institutional review board (IRB) protocol #IRB00249128 hosted at Johns Hopkins University School of Medicine via the SMART IRB40 Master Common Reciprocal reliance agreement. This central IRB cover data contributions and transfer to N3C and does not cover research using N3C data. If elected, individual sites may choose to exercise their own local IRB agreements instead of utilizing the central IRB. As NCATs is the steward of the repository, data received and hosted by NCATs on the N3C Data Enclave, its maintenance, and its storage are covered under a central NIH IRB protocol to make EHR-derived data available for the clinical and research community to use for studying COVID-19. Our institution has an active data transfer agreement with N3C. This specific analysis of the N3C Enclave was approved by N3C under the Data Use Agreement titled “[RP-7C5E62] COVID-19 Outcomes in Patients with Cirrhosis.” The use of N3C data for this study was authorized by the IRB at the University of California, San Francisco under #20-33149.

## Results

As of May 15, 2021, 56 sites that had completed data transfer were harmonized and integrated into the N3C Enclave. This included approximately 6.6 billion rows of data on 5,904,987 unique patients, of which 4,786,983 patients had at least one SARS-CoV-2 test per above definitions. Of these approximately 4.8 million patients who had undergone testing, an analytical sample of 217,143 CLD patients with or without cirrhosis was assembled, after applying above exclusion criteria for transplant status, age, and accounting for date shifting in the N3C Enclave (Figure 1). Based on SARS-CoV-2 test results, we divided the 217,143 CLD patients into the four cohorts: 129,097 (59%) Non-Cirrhosis/Negative, 25,844 (12%) Non-Cirrhosis/Positive, 54,065 (25%) Cirrhosis/Negative, and 8,137 (4%) Cirrhosis/Positive.

### Demographic and Clinical Characteristics

The baseline demographic and clinical characteristics of the four cohorts are presented in Table 1. In general, the four cohorts differed significantly with regards to distributions of age, race/ethnicity, etiologies of chronic liver disease, CCI scores, NREVSS regions, and laboratory test values. Of note, patients with cirrhosis in our cohorts were less likely to be women: 52% of Non-Cirrhosis/Negative, 53% of Non-Cirrhosis/Positive, and 44% of Cirrhosis/Negative and 43% of Cirrhosis/Positive cohorts were women. Of CLD etiologies, there were notably differences in the distribution of patients with AALD: 34% and 29% of the Cirrhosis/Negative and Cirrhosis/Positive cohorts, respectively, compared to 6% and 7% of the Non-Cirrhosis/Negative and Non-Cirrhosis/Positive cohorts, respectively.

**Table 1.**
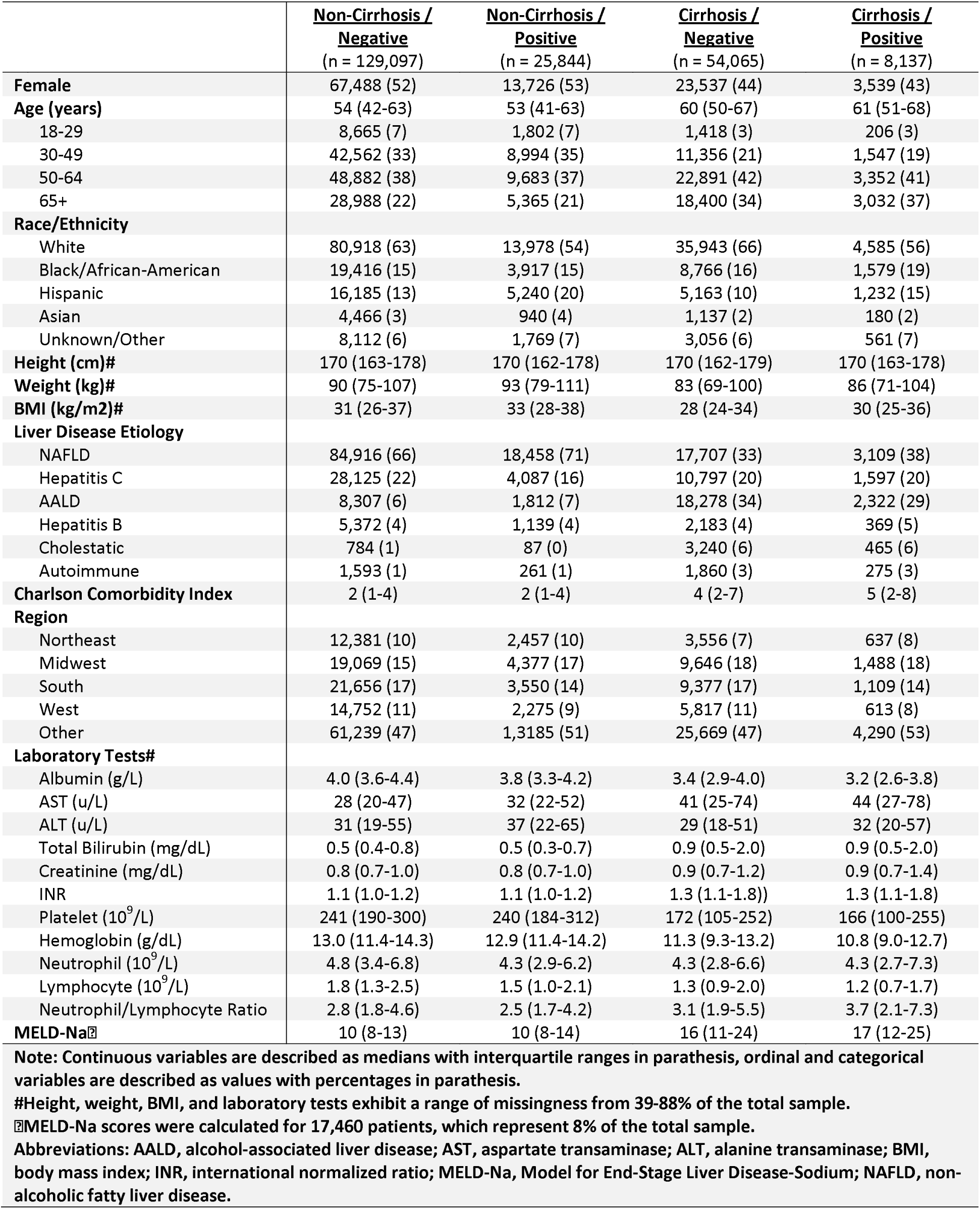
Baseline demographic, clinical, and laboratory characteristics of the 217,143 patients with chronic liver diseases with and without cirrhosis included in the analysis.

|  | <u>Non-Cirrhosis /</u><br><u>Negative</u><br>(n = 129,097) | <u>Non-Cirrhosis /</u><br><u>Positive</u><br>(n = 25,844) | <u>Cirrhosis /</u><br><u>Negative</u><br>(n = 54,065) | <u>Cirrhosis /</u><br><u>Positive</u><br>(n = 8,137) |
| --- | --- | --- | --- | --- |
| **Female** | 67,488 (52) | 13,726 (53) | 23,537 (44) | 3,539 (43) |
| **Age (years)** | 54 (42-63) | 53 (41-63) | 60 (50-67) | 61 (51-68) |
| 18-29 | 8,665 (7) | 1,802 (7) | 1,418 (3) | 206 (3) |
| 30-49 | 42,562 (33) | 8,994 (35) | 11,356 (21) | 1,547 (19) |
| 50-64 | 48,882 (38) | 9,683 (37) | 22,891 (42) | 3,352 (41) |
| 65+ | 28,988 (22) | 5,365 (21) | 18,400 (34) | 3,032 (37) |
| **Race/Ethnicity** |  |  |  |  |
| White | 80,918 (63) | 13,978 (54) | 35,943 (66) | 4,585 (56) |
| Black/African-American | 19,416 (15) | 3,917 (15) | 8,766 (16) | 1,579 (19) |
| Hispanic | 16,185 (13) | 5,240 (20) | 5,163 (10) | 1,232 (15) |
| Asian | 4,466 (3) | 940 (4) | 1,137 (2) | 180 (2) |
| Unknown/Other | 8,112 (6) | 1,769 (7) | 3,056 (6) | 561 (7) |
| **Height (cm)#** | 170 (163-178) | 170 (162-178) | 170 (162-179) | 170 (163-178) |
| **Weight (kg)#** | 90 (75-107) | 93 (79-111) | 83 (69-100) | 86 (71-104) |
| **BMI (kg/m2)#** | 31 (26-37) | 33 (28-38) | 28 (24-34) | 30 (25-36) |
| **Liver Disease Etiology** |  |  |  |  |
| NAFLD | 84,916 (66) | 18,458 (71) | 17,707 (33) | 3,109 (38) |
| Hepatitis C | 28,125 (22) | 4,087 (16) | 10,797 (20) | 1,597 (20) |
| AALD | 8,307 (6) | 1,812 (7) | 18,278 (34) | 2,322 (29) |
| Hepatitis B | 5,372 (4) | 1,139 (4) | 2,183 (4) | 369 (5) |
| Cholestatic | 784 (1) | 87 (0) | 3,240 (6) | 465 (6) |
| Autoimmune | 1,593 (1) | 261 (1) | 1,860 (3) | 275 (3) |
| **Charlson Comorbidity Index** | 2 (1-4) | 2 (1-4) | 4 (2-7) | 5 (2-8) |
| **Region** |  |  |  |  |
| Northeast | 12,381 (10) | 2,457 (10) | 3,556 (7) | 637 (8) |
| Midwest | 19,069 (15) | 4,377 (17) | 9,646 (18) | 1,488 (18) |
| South | 21,656 (17) | 3,550 (14) | 9,377 (17) | 1,109 (14) |
| West | 14,752 (11) | 2,275 (9) | 5,817 (11) | 613 (8) |
| Other | 61,239 (47) | 1,3185 (51) | 25,669 (47) | 4,290 (53) |
| **Laboratory Tests#** |  |  |  |  |
| Albumin (g/L) | 4.0 (3.6-4.4) | 3.8 (3.3-4.2) | 3.4 (2.9-4.0) | 3.2 (2.6-3.8) |
| AST (u/L) | 28 (20-47) | 32 (22-52) | 41 (25-74) | 44 (27-78) |
| ALT (u/L) | 31 (19-55) | 37 (22-65) | 29 (18-51) | 32 (20-57) |
| Total Bilirubin (mg/dL) | 0.5 (0.4-0.8) | 0.5 (0.3-0.7) | 0.9 (0.5-2.0) | 0.9 (0.5-2.0) |
| Creatinine (mg/dL) | 0.8 (0.7-1.0) | 0.8 (0.7-1.0) | 0.9 (0.7-1.2) | 0.9 (0.7-1.4) |
| INR | 1.1 (1.0-1.2) | 1.1 (1.0-1.2) | 1.3 (1.1-1.8) | 1.3 (1.1-1.8) |
| Platelet (10<sup>9</sup>/L) | 241 (190-300) | 240 (184-312) | 172 (105-252) | 166 (100-255) |
| Hemoglobin (g/dL) | 13.0 (11.4-14.3) | 12.9 (11.4-14.2) | 11.3 (9.3-13.2) | 10.8 (9.0-12.7) |
| Neutrophil (10<sup>9</sup>/L) | 4.8 (3.4-6.8) | 4.3 (2.9-6.2) | 4.3 (2.8-6.6) | 4.3 (2.7-7.3) |
| Lymphocyte (10<sup>9</sup>/L) | 1.8 (1.3-2.5) | 1.5 (1.0-2.1) | 1.3 (0.9-2.0) | 1.2 (0.7-1.7) |
| Neutrophil/Lymphocyte Ratio | 2.8 (1.8-4.6) | 2.5 (1.7-4.2) | 3.1 (1.9-5.5) | 3.7 (2.1-7.3) |
| **MELD-Na** | 10 (8-13) | 10 (8-14) | 16 (11-24) | 17 (12-25) |

**Note:** Continuous variables are described as medians with interquartile ranges in parathesis, ordinal and categorical variables are described as values with percentages in parathesis.
**#**Height, weight, BMI, and laboratory tests exhibit a range of missingness from 39-88% of the total sample.
**▣**MELD-Na scores were calculated for 17,460 patients, which represent 8% of the total sample.
**Abbreviations:** AALD, alcohol-associated liver disease; AST, aspartate transaminase; ALT, alanine transaminase; BMI, body mass index; INR, international normalized ratio; MELD-Na, Model for End-Stage Liver Disease-Sodium; NAFLD, non-alcoholic fatty liver disease.

For the 17,460 (8%) patients where all MELD-Na components were available and scores were calculated, the median (IQR) MELD-Na were 16 (11-24) and 17 (12-25) in Cirrhosis/Negative and Cirrhosis/Positive patients, respectively, compared to 10 (8-13) and 10 (8-14) in Non-Cirrhosis/Negative and Non-Cirrhosis/Positive patients. For 112,760 (52%) CLD patients whose location data were available, we found that patients were represented from every CDC NREVVS region and state except for North Dakota in both in the full sample and among those patients with positive SARS-CoV-2 tests (Figure 2).

**Figure 2.**
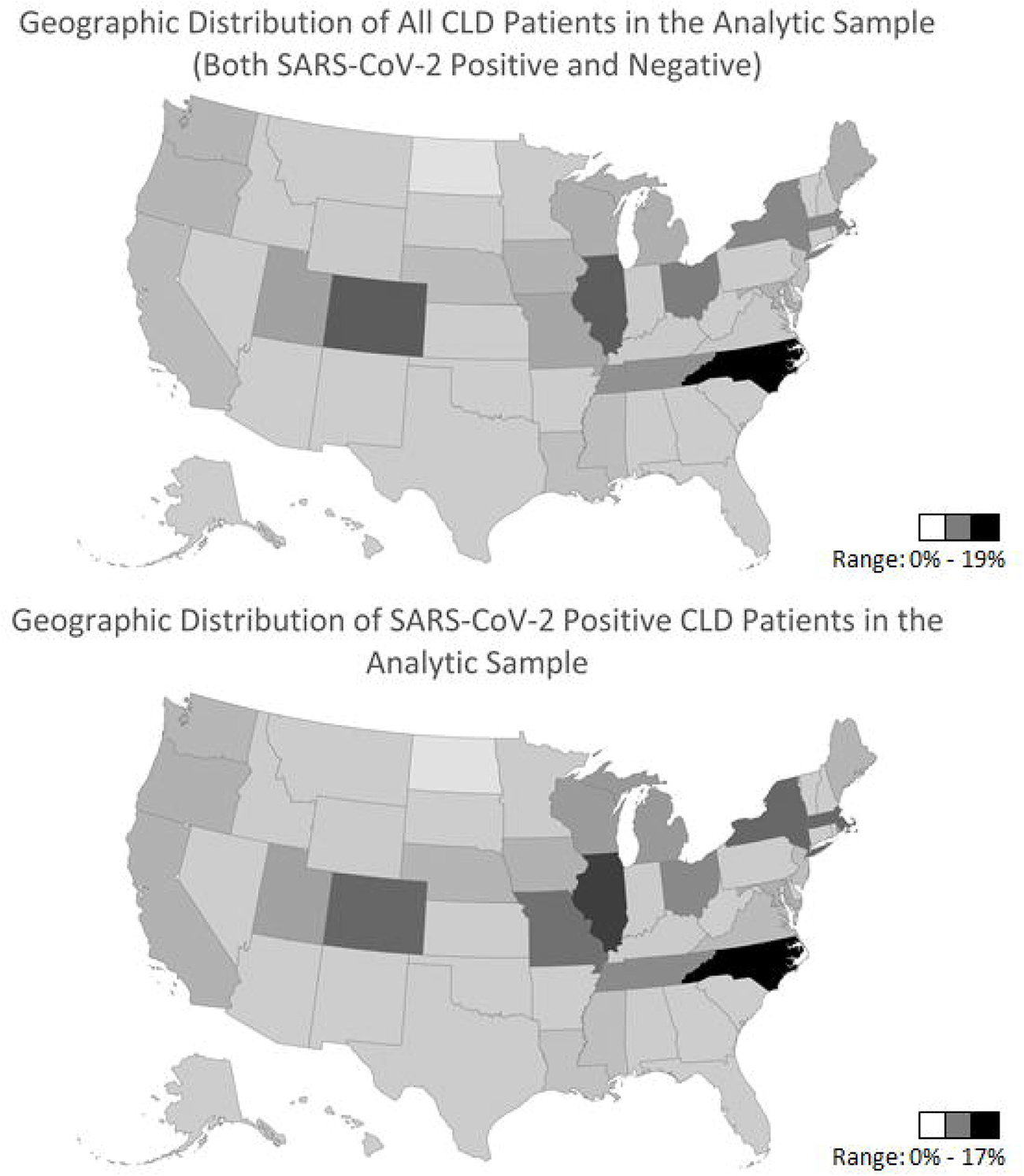
Geographic distributions of CLD patients and CLD patients with positive SARS-CoV-2 testing in analytic sample.

### Death, Hospitalization, and Mechanical Ventilation Rates

Cumulative incidences of outcomes of interest, including 30- and 90-day all-cause death, hospitalization, and mechanical ventilation, are presented in Table 2. Thirty-day all-cause death rates increased progressively from 0.4% in Non-Cirrhosis/Negative patients to 1.8% in Non-Cirrhosis/Positive patients, and from 4.0% in Cirrhosis/Negative patients to 9.7% in Cirrhosis/Positive patients. Ninety-day all-cause death rates also increased progressively from 0.8% in Non-Cirrhosis/Negative patients to 2.3% in Non-Cirrhosis/Positive patients, and from 7.3% in Cirrhosis/Negative patients to 14.0% in Cirrhosis/Positive patients. Thirty- and 90-day mechanical ventilation rates also increased in a similar fashion based on SARS-CoV-2 status and presence of cirrhosis (Table 2).

**Table 2.** Cumulative incidences of mortality, mechanical ventilation, and hospitalization at 30- and 90-days after the index date.

| Cumulative Incidence Rates (95%CI) | <u>Non-Cirrrosis /</u><br><u>Negative</u><br>(n = 129,097) | <u>Non-Cirrrosis /</u><br><u>Positive</u><br>(n = 25,844) | <u>Cirrrosis /</u><br><u>Negative</u><br>(n = 54,065) | <u>Cirrrosis /</u><br><u>Positive</u><br>(n = 8,137) |
| --- | --- | --- | --- | --- |
| Hospitalization by Day 30 | 27.5 (28.8-27.2) | 21.4 (18.7-20.9) | 48.7 (49.9-48.3) | 48.3 (44.6-47.2) |
| Hospitalization by Day 90 | 30 (29.7-30.2) | 24.1 (23.5-24.6) | 51.8 (51.4-52.2) | 51.8 (50.6-52.9) |
| Mechanical Ventilation by Day 30 | 0.9 (0.8-0.9) | 2.2 (2.1-2.4) | 5.4 (5.2-5.6) | 10.4 (9.8-11.1) |
| Mechanical Ventilation by Day 90 | 1.1 (1.0-1.1) | 2.4 (2.2-2.6) | 6.8 (6.6-7.0) | 11.8 (11.1-12.5) |
| Mortality by Day 30 | 0.4 (0.4-0.4) | 1.8 (1.6-2.0) | 4.0 (3.9-4.2) | 9.7 (9.0-10.4) |
| Mortality by Day 90 | 0.8 (0.8-0.9) | 2.3 (2.1-2.5) | 7.3 (7.0-7.5) | 14.0 (13.2-14.8) |

**Note:** Cumulative incidences are expressed as percentages. **Abbreviations:** CI, confidence interval.

Of note, 30-day and 90-day hospitalization rates were consistently higher among patients with cirrhosis (both Cirrhosis/Negative and Cirrhosis/Positive) compared to those patients without cirrhosis (both Non-Cirrhosis/Negative and non-Cirrhosis/Positive). Among both patients with and without cirrhosis, those testing negative for SARS-CoV-2 (Non-Cirrhosis/Negative and Cirrhosis/Negative) had higher 30- and 90-day hospitalization rates. Kaplan-Meier curves for 30-day mortality among the four cohorts are presented in Figure 3.

**Figure 3.**
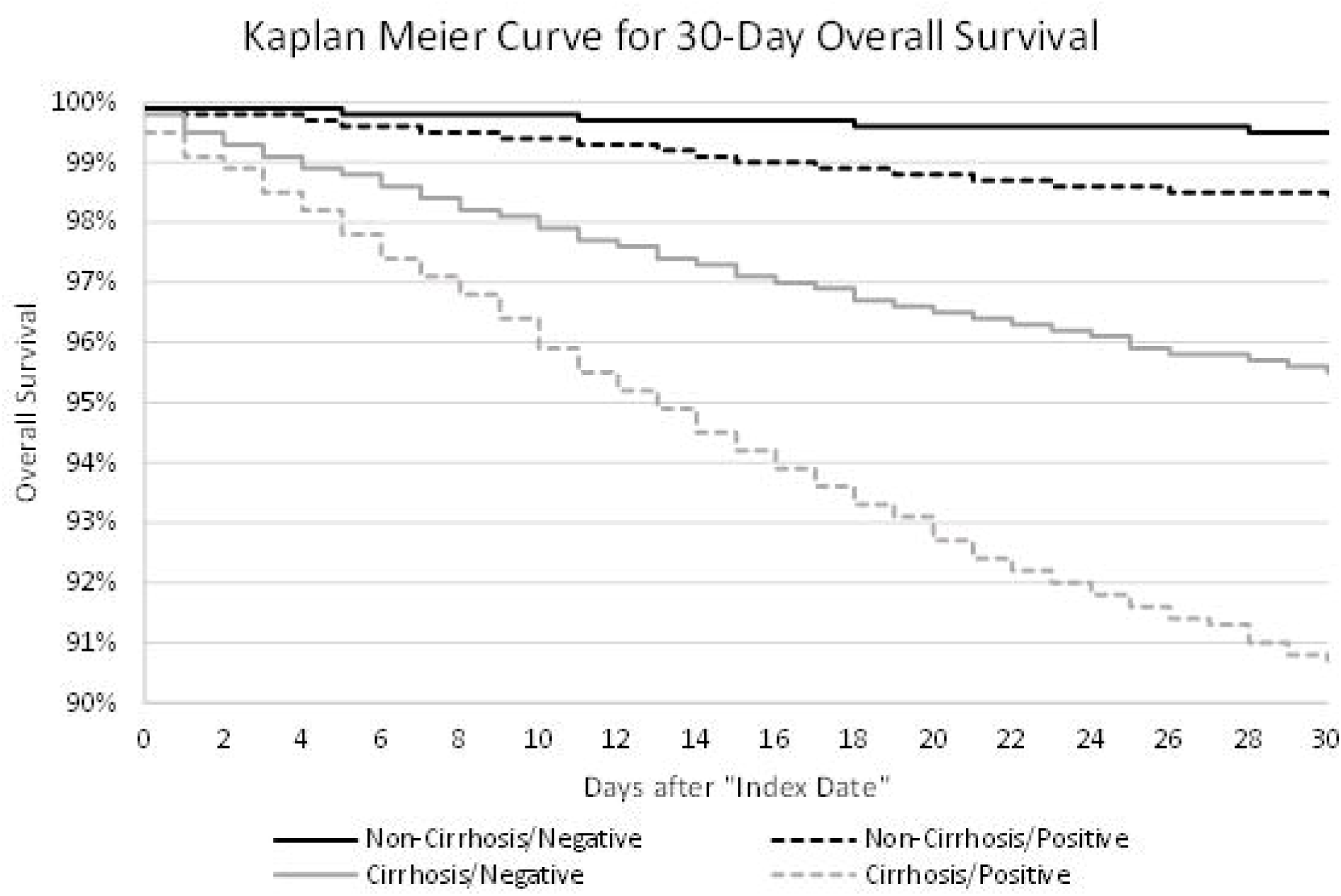
Kaplan-Meier Curves for 30-day Survival of CLD Patients in the Analytic Sample.

### Association between SARS-CoV-2 Infection and Death in Patients with Cirrhosis

In univariate analyses, compared to Cirrhosis/Negative patients, Cirrhosis/Positive patients was associated with 2.5-times hazard of death within 30 days (hazard ratio [HR] 2.49, 95% confidence interval [CI] 2.29-2.71, p<0.01). In multivariate analyses, compared to Cirrhosis/Negative patients, Cirrhosis/Positive patients was associated with 2.4-times hazard of death within 30-days (adjusted hazard ratio [aHR] 2.43, 95%CI 2.23-2.64, p<0.01) after adjusting for race/ethnicity, CLD etiology, CCI index, and region.

Of note, age (aHR 1.02, 95%CI 1.01-1.02, p<0.01), Other/Unknown race/ethnicity (aHR 1.37, 95%CI 1.18-1.59, p<0.01), AALD as etiology (aHR 1.34, 95%CI 1.22-1.47, p<0.01), and Charlson Comorbidity Index (aHR 1.07 per point, 95%CI 1.06-1.08, p<0.01) were associated with higher 30-day mortality hazards in multivariate analyses. Location in Other/Unknown region (aHR 0.73, 95%CI 0.63-0.84, p<0.01) were associated with lower 30-day mortality hazards in multivariate analyses. Detailed results are presented in Table 3.

**Table 3.** Association of SARS-CoV-2 infection with all-cause 30-day mortality in patients with cirrhosis (Cirrhosis/Positive versus Cirrhosis/Negative)

|  | Univariable Cox Regression |  |  | Multivariable Cox Regression |  |  |
| --- | --- | --- | --- | --- | --- | --- |
|  | HR | 95% CI | P-Value | aHR | 95% CI | P-Value |
| SARS-CoV-2 Infection | 2.49 | 2.29-2.71 | <0.01 | 2.43 | 2.23-2.64 | <0.01 |
| Age (years) | 1.02 | 1.02-1.02 | <0.01 | 1.02 | 1.01-1.02 | <0.01 |
| Female | 0.89 | 0.82-0.96 | <0.01 | 0.96 | 0.89-1.04 | 0.30 |
| Race/Ethnicity, no. (%) |  |  |  |  |  |  |
| White | Ref |  |  | Ref |  |  |
| Black/African-American | 1.13 | 1.02-1.25 | 0.01 | 1.04 | 0.93-1.15 | 0.51 |
| Hispanic | 1.11 | 0.98-1.25 | 0.09 | 1.07 | 0.95-1.21 | 0.27 |
| Asian | 0.86 | 0.65-1.14 | 0.30 | 0.88 | 0.66-1.17 | 0.38 |
| Unknown/Other | 1.27 | 1.10-1.48 | <0.01 | 1.37 | 1.18-1.59 | <0.01 |
| Etiology of Liver Disease, no. (%) |  |  |  |  |  |  |
| NAFLD | Ref |  |  | Ref |  |  |
| Hepatitis C | 0.89 | 0.80-1.00 | 0.05 | 0.93 | 0.83-1.04 | 0.22 |
| AALD | 1.17 | 1.07-1.27 | <0.01 | 1.34 | 1.22-1.47 | <0.01 |
| Hepatitis B | 0.96 | 0.78-1.16 | 0.65 | 0.97 | 0.79-1.18 | 0.74 |
| Cholestatic | 0.58 | 0.47-0.72 | <0.01 | 0.64 | 0.52-0.79 | <0.01 |
| Autoimmune | 0.82 | 0.65-1.03 | 0.08 | 0.89 | 0.71-1.12 | 0.31 |
| Charlson Comorbidity Index | 1.08 | 1.07-1.09 | <0.01 | 1.07 | 1.06-1.08 | <0.01 |
| Region |  |  |  |  |  |  |
| Northeast | Ref |  |  | Ref |  |  |
| Midwest | 0.92 | 0.79-1.08 | 0.32 | 0.91 | 0.78-1.07 | 0.26 |
| South | 0.98 | 0.84-1.15 | 0.80 | 1.03 | 0.88-1.21 | 0.73 |
| West | 0.82 | 0.69-0.98 | 0.03 | 0.88 | 0.74-1.05 | 0.17 |
| Other | 0.75 | 0.65-0.87 | <0.01 | 0.73 | 0.63-0.84 | <0.01 |

**Abbreviations:** AALD, alcohol-associated liver disease; aHR, adjusted hazard ratio; CI, confidence interval; HR, hazard ratio; NAFLD, non-alcoholic fatty liver disease.

### Association between Presence of Cirrhosis and Death in CLD Patients who tested SARS-CoV-2 Positive

In univariate analyses, compared to Non-Cirrhosis/Positive patients, the presence of cirrhosis (Cirrhosis/Positive) was associated with 5.6-times hazard of death within 30 days (HR 5.57, 95%CI 54.94-6.29, p<0.01). In multivariate analyses, compared to Non-Cirrhosis/Positive patients, the presence of cirrhosis (Cirrhosis/Positive) was associated with a 3.4-times hazard of death within 30 days (aHR 3.39, 95%CI 2.96-3.89, p<0.01) after adjusting for race/ethnicity, CLD etiology, CCI index, and region.

Of note, age (aHR 1.05 per year, 95%CI 1.05-1.06, p<0.01), Hispanic ethnicity (aHR 1.33, 95%CI 1.13-1.57, p<0.01), Other/Unknown race (aHR 1.61, 95%CI 1.29-2.00, p<0.01), AALD (aHR 1.28, 95%CI 1.08-1.41, p<0.01), and Charlson Comorbidity Index (aHR 1.06 per point, 95%CI 1.05-1.08, p<0.01) were associated with higher 30-day mortality hazards in multivariate analyses. Female gender (aHR 0.84, 95%CI 0.74-0.95, p<0.01), location in the Midwest (aHR 0.58, 95%CI 0.47-0.72, p<0.01), location in the West (aHR 0.54, 95%CI 0.40-0.72, p<0.01) and Other/Unknown locations (aHR 0.55, 95%CI 0.46-0.66, p<0.01) were associated with lower 30-day mortality hazards in multivariate analyses. Detailed results are presented in Table 4.

**Table 4.** Association of presence of cirrhosis with all-cause 30-day mortality in all CLD patients who tested positive for SARS-CoV-2 infection (Cirrhosis/Positive versus Non-Cirrhosis/Positive)

|  | Univariable Cox Regression |  |  | Multivariable Cox Regression |  |  |
| --- | --- | --- | --- | --- | --- | --- |
|  | HR | 95% CI | P-Value | aHR | 95% CI | P-Value |
| Presence of Cirrhosis | 5.57 | 4.94-6.29 | <0.01 | 3.39 | 2.96-3.89 | <0.01 |
| Age (years) | 1.07 | 1.06-1.07 | <0.01 | 1.05 | 1.05-1.06 | <0.01 |
| Female | 0.65 | 0.58-0.73 | <0.01 | 0.84 | 0.74-0.95 | <0.01 |
| Race/Ethnicity |  |  |  |  |  |  |
| White | Ref |  |  | Ref |  |  |
| Black/African-American | 1.29 | 1.11-1.51 | <0.01 | 1.06 | 0.90-1.24 | 0.52 |
| Hispanic | 0.90 | 0.77-1.07 | 0.23 | 1.33 | 1.13-1.57 | <0.01 |
| Asian | 0.97 | 0.69-1.36 | 0.85 | 1.28 | 0.90-1.83 | 0.16 |
| Unknown/Other | 1.38 | 1.11-1.71 | <0.01 | 1.61 | 1.29-2.00 | <0.01 |
| Etiology of Liver Disease |  |  |  |  |  |  |
| NAFLD | Ref |  |  | Ref |  |  |
| Hepatitis C | 1.86 | 1.60-2.16 | <0.01 | 1.22 | 1.04-1.43 | 0.02 |
| AALD | 2.50 | 2.15-2.90 | <0.01 | 1.28 | 1.08-1.51 | <0.01 |
| Hepatitis B | 1.43 | 1.08-1.90 | 0.01 | 1.00 | 0.75-1.34 | 0.99 |
| Cholestatic | 1.98 | 1.34-2.91 | <0.01 | 0.73 | 0.50-1.09 | 0.12 |
| Autoimmune | 2.05 | 1.40-2.99 | <0.01 | 1.12 | 0.76-1.64 | 0.58 |
| Charlson Comorbidity Index | 1.18 | 1.16-1.19 | <0.01 | 1.06 | 1.05-1.08 | <0.01 |
| Region |  |  |  |  |  |  |
| Northeast | Ref |  |  | Ref |  |  |
| Midwest | 0.64 | 0.51-0.79 | <0.01 | 0.58 | 0.47-0.72 | <0.01 |
| South | 0.86 | 0.70-1.06 | 0.15 | 0.87 | 0.70-1.07 | 0.19 |
| West | 0.45 | 0.34-0.60 | <0.01 | 0.54 | 0.40-0.72 | <0.01 |
| Other | 0.59 | 0.50-0.71 | <0.01 | 0.55 | 0.46-0.66 | <0.01 |
| Abbreviations: AALD, alcohol-associated liver disease; aHR, adjusted hazard ratio; CI, confidence interval; HR, hazard ratio; NAFLD, non-alcoholic fatty liver disease. |  |  |  |  |  |  |

### Factors Associated with 30-Day Mortality among Cirrhosis/Positive Patients

Demographic and clinical factors associated with all-cause 30-day mortality among Cirrhosis/Positive patients are presented in Table 5. In univariate analyses, we found that age (HR 1.04 per year, 95%CI 1.03-1.04, p<0.01), Other/Unknown race (HR 1.47, 95%CI 1.13-1.91, p<0.01), and Charlson Comorbidity Index (HR 1.06, 95%CI 1.04-1.08, p<0.01) were associated with higher risk of 30-day mortality among Cirrhosis/Positive patients. Cholestatic liver diseases (HR 0.62, 95%CI 0.43-0.92, p=0.02), location in the West (HR 0.66, 95%CI 0.47-0.93, p=0.02), and Other/Unknown locations (HR 0.57, 95%CI 0.45-0.73, p<0.01) were associated with lower hazards of mortality in univariate analyses.

**Table 5.** Factors associated with all-cause 30-day mortality among patients with cirrhosis who tested positive for SARS-CoV-2 infection (Cirrhosis/Positive patients only)

|  | Univariable Cox Regression |  |  | Multivariable Cox Regression |  |  |
| --- | --- | --- | --- | --- | --- | --- |
|  | HR | 95% CI | P-Value | aHR | 95% CI | P-Value |
| **Age (years)** | 1.04 | 1.03-1.04 | <0.01 | 1.03 | 1.03-1.04 | <0.01 |
| **Female** | 0.97 | 0.84-1.13 | 0.72 | 1.02 | 0.87-1.19 | 0.80 |
| **Race/Ethnicity** |  |  |  |  |  |  |
| White | Ref |  |  | Ref |  |  |
| Black/African-American | 1.05 | 0.87-1.27 | 0.63 | 1.06 | 0.87-1.30 | 0.57 |
| Hispanic | 1.11 | 0.90-1.37 | 0.32 | 1.24 | 1.00-1.53 | 0.05 |
| Asian | 1.25 | 0.79-1.98 | 0.34 | 1.26 | 0.78-2.02 | 0.34 |
| Unknown/Other | 1.47 | 1.13-1.91 | <0.01 | 1.65 | 1.27-2.15 | <0.01 |
| **Etiology of Liver Disease** |  |  |  |  |  |  |
| NAFLD | Ref |  |  | Ref |  |  |
| Hepatitis C | 0.93 | 0.76-1.14 | 0.46 | 0.91 | 0.73-1.12 | 0.37 |
| AALD | 1.01 | 0.85-1.20 | 0.91 | 1.11 | 0.92-1.34 | 0.27 |
| Hepatitis B | 0.95 | 0.66-1.36 | 0.76 | 0.88 | 0.61-1.28 | 0.52 |
| Cholestatic | 0.62 | 0.42-0.92 | 0.02 | 0.63 | 0.42-0.94 | 0.02 |
| Autoimmune | 0.93 | 0.61-1.41 | 0.73 | 0.98 | 0.65-1.50 | 0.94 |
| **Charlson Comorbidity Index** | 1.06 | 1.04-1.08 | <0.01 | 1.04 | 1.02-1.06 | <0.01 |
| **Region** |  |  |  |  |  |  |
| Northeast | Ref |  |  | Ref |  |  |
| Midwest | 0.58 | 0.44-0.77 | <0.01 | 0.60 | 0.45-0.80 | <0.01 |
| South | 0.77 | 0.58-1.02 | 0.07 | 0.83 | 0.62-1.11 | 0.21 |
| West | 0.66 | 0.47-0.93 | 0.02 | 0.75 | 0.54-1.05 | 0.10 |
| Other | 0.57 | 0.45-0.73 | <0.01 | 0.58 | 0.46-0.74 | <0.01 |

Abbreviations: AALD, alcohol-associated liver disease; aHR, adjusted hazard ratio; CI, confidence interval; HR, hazard ratio; NAFLD, non-alcoholic fatty liver disease.

In multivariate analyses, age (aHR 1.03 per year, 95%CI 1.03-1.04, p<0.01), Hispanic ethnicity (aHR 1.24, 95%CI 1.00-1.53, p=0.05), Other/Unknown race (aHR 1.65, 95%CI 1.27-2.15, p<0.01) and Charlson Comorbidity Index (aHR 1.04, 95%CI 1.02-1.06, p<0.01) were associated with higher hazards of 30-day mortality. Cholestatic liver diseases (aHR 0.63, 95%CI 0.42-0.94, p<0.01), location in the Midwest (aHR 0.60, 95%CI 0.45-0.80, p<0.01) and Other/Unknown location (aHR 0.58, 95%CI 0.46-0.74, p<0.01)were associated with lower hazards of 30-day mortality in multivariate analyses.

### Sensitivity Analyses with MELD-Na and Serum Albumin

As calculated MELD-Na scores were available for only 17,460 (8%) patients and serum albumin values were available for 73,313 (34%) patients, we conducted sensitivity analyses to determine the influence of these variables on the above multivariate models comparing patients with cirrhosis (Supplemental Table 4). For the multivariate model evaluating the association of SARS-CoV-2 infection with death in patients with cirrhosis, further adjustments for MELD-Na and serum albumin did not change the significance of the association (aHR ranged from 1.81 to 2.43). For the multivariate model evaluating factors associated with death among Cirrhosis/Positive patients, further adjustments for MELD-Na and serum albumin did not change the significance of the association for age and death (aHR ranged from 1.02 to 1.04). These adjustments for MELD-Na and serum albumin did, however, eliminate the associations of race/ethnicity (Hispanic and Other/Unknown) and Charlson Comorbidity Index with increased hazard of death. Similarly, these adjustments eliminate the associations of CLD etiology (cholestatic liver disease) and location (Midwest and Other/Unknown) with decreased hazards of death.

## Discussion

In this study of nearly 220,000 patients with chronic liver disease in the National COVID Cohort Collaborative, we found that SARS-CoV-2 infection was associated with greater than two-fold hazard of all-cause mortality within 30 days among patients with cirrhosis. Among all chronic liver disease patients (with and without cirrhosis) who tested SARS-CoV-2 positive, the presence of cirrhosis was associated with three-fold hazard of all-cause mortality within 30 days.

Our results are largely consistent with previous studies of chronic liver disease patients with and without cirrhosis who tested positive for SARS-CoV-2 infection. Our use of the N3C Data Enclave, haver, has several unique features that enhance the generalizability of our results and advance our understanding of COVID-19 in CLD patients. The N3C Data Enclave harmonizes and unifies data from a very large number of sites (208 have signed data transfer agreements as of May 15, 2021).^13^ As site-to-site variations in demographic, diagnostic, and outcome measurements have been previously reported, the use of a centralized curation process ensures quality across disparate data between sites. Moreover, all data available on the N3C Data Enclave have been synchronized into the OMOP common data model, thereby allowing reproducible queries, transformations, and analyses executed on other OMOP-based databases.^15,16,30^

The number of clinical sites included in this specific study (harmonized data from 56 sites as of May 15, 2021) confers a major strength to this study in terms of the number of patients (nearly 160,000), national scope, and demographic representation. Notably, 50% of the participants included in our study were women and 31% were racial/ethnic minorities: 16% identified as Black/African-American, 13% Hispanic, and 3% Asian. Consistent with previous studies of COVID-19 in the general population, we found female gender was associated with a lower hazard of death (aHR 0.84, 95%CI 0.74-0.95, p<0.01) among all CLD patients with SARS-CoV-2 infection (Table 4). This gender association, however, did not remain once we stratified to only Cirrhosis/Positive patients to seek out factors associated with mortality in this specific population.

Consistent with extensive racial/ethnic disparities described in COVID-19 related outcomes,^31,32^ we found an increased hazard of mortality for those who identified as Hispanic (aHR 1.33, 95%CI 1.13-1.57, p<0.01) and those who identified as Other/Unknown (aHR 1.61, 95%CI 1.29-2.00, p<0.01) among CLD patients with positive SARS-CoV-2 test (Table 4). These results likely reflect the broader racial/ethnic disparities concerning differential rates of SARS-CoV-2 infection and socioeconomic and longstanding structural disparities in access to care. The research questions regarding racial/ethnic disparities concerning differential rates of SARS-CoV-2 infection is an active area of exploration of several N3C research teams.^12–14^

Compared to previous studies which included data in the early phases of the COVID-19 pandemic, the data from our study covers a longer duration up to May 2021 and reflects changes in treatment algorithms and therapy advances. For example, we found a lower cumulative incidence for all-cause mortality at 30-days at 9.7% for Cirrhosis/Positive patients compared to previous studies with estimates up to 17%.^11^ We also found that the 30- and 90-day hospitalization rates were higher in the SARS-CoV-2 negative groups (Non-Cirrhosis/negative and Cirrhosis/negative patients). This is likely due to changes in healthcare delivery during the COVID-19 pandemic as standardized asymptomatic testing prior to hospital admissions and procedures became widespread.^33,34^ As such, the Non-Cirrhosis/Negative and Cirrhosis/Negative populations were subject to selection bias as these cohorts were more likely to undergo procedures or be hospitalized than the general population of CLD patients.

We acknowledge the following limitations. First, as data were aggregated from many sites, there is systematic missingness of certain variables. In our study, this is most apparent in that we were only able to calculate the MELD-Na scores for 17,460 patients. We accounted for this by conducting sensitivity analyses that showed our main finding of SARS-CoV-2 infection in patients with cirrhosis was associated with two-fold hazard in mortality did not change. In addition, our sensitivity analyses revealed that certain geographic and CLD etiology associations with mortality were eliminated once adjustments for MELD-Na and serum albumin were made in Cirrhosis/Positive patients. This most likely reflected that data missingness in N3C occurs not at random and is associated with certain sites. Second, although N3C has standardized protocols for data curation and harmonization, there likely remains variations in terminology and ontology between various sites. The use of the OMOP common data model, however, decreases such differences and enforces a degree of standardization.^15,16,30^

Third, due to date-shifting employed in the process of de-identification in the N3C Data Enclave and differences in data harmonization times between data partner sites, there may be a delay in ascertainment of outcomes. This means that there may have been misclassification of outcomes if the date of SARS-CoV-2 testing was close to the latest known date of records (“maximum data date”) for that data partner site – e.g. the outcome has not been recorded and harmonized into the N3C Data Enclave. To account for these issues, we employed two methods: 1. We attempted to maximize follow up for each patient by defining last follow up as any encounters or records (visit occurrence, procedure, measurement, observation, or condition occurrence) in the OMOP data model instead of strictly based clinic or hospital visits. 2. We excluded patients whose date of SARS-CoV-2 testing was within 90 days of the maximum data date – this exclusion criteria only affected 1% of potential patients to be included in the analytical sample.

Fourth, we utilized the de-identified version of the N3C Data Enclave to conduct our analyses. To protect patient privacy, date shifting uniformly was applied to all data. This means that our analyses could not investigate temporal trends with each COVID-19 surge in the United States. Lastly, there is likely misclassification between patients with AALD and NAFLD given the non-specific nature of OMOP concept identifier 4059290 “steatosis of the liver” (corresponding to ICD-10 code K76.0). This is most apparent in only 7% of CLD patients without cirrhosis were classified to have AALD while 33% of patients with cirrhosis were classified with AALD (due to more specific ICD-10 codes for cirrhosis due to AALD).

Despite these limitations, our study is one of the largest studies of outcomes of SARS-CoV-2 infection in patients with chronic liver diseases with and without cirrhosis to date. In addition, this is one of the first studies within gastroenterology and hepatology that specifically utilized the OMOP common data model for data extraction and primary analyses. Given the continued expansion of N3C and ongoing acquisition of longitudinal data, our study in the N3C Data Enclave lays the foundation for studying future potential clinical questions affecting gastroenterology and hepatology patient populations as the COVID-19 pandemic continues to evolve.^35^

## Data Availability

The N3C Data Enclave (covid.cd2h.org/enclave) houses fully reproducible, transparent, and broadly available limited and de-identified datasets (HIPAA definitions: https://www.hhs.gov/hipaa/for-professionals/privacy/specialtopics/de-identification/index.html). Data is accessible by investigators at institutions that have signed a Data Use Agreement with NIH who have taken human subjects and security training and attest to the N3C User Code of Conduct. Investigators wishing to access the limited dataset must also supply an institutional IRB protocol. All requests for data access are reviewed by the NIH Data Access Committee. A full description of the N3C Enclave governance has been published;193 information about how to apply for access is available on the NCATS website: https://ncats.nih.gov/n3c/about/applying-for-access. Reviewers and health authorities will be given access permission and guidance to aid reproducibility and outcomes assessment. A Frequently Asked Questions about the data and access has been created at: https://ncats.nih.gov/n3c/about/program-faq The data model is OMOP 5.3.1, specifications are posted at: https://ncats.nih.gov/files/OMOP_CDM_COVID.pdf

## Abbreviations

AALD: alcohol-associated liver disease
aHR: adjusted hazard ratio
AST: aspartate transaminase
ALT: alanine transaminase
BMI: body mass index
CCI: Charlson Comorbidity Index
CD2H: Center for Data to Health
CDC: Centers for Disease Control and Prevention
CLD: chronic liver disease
CTSA: Clinical and Translational Science Awards
EHR: electronic health record
ICD-10-CM: International Classification of Diseases, Tenth Revision, Clinical Modification
HR: hazard ratio
INR: international normalized ratio
IQR: interquartile range
IRB: institutional review board
MELD-Na: Model for End-Stage Liver Disease-Sodium
N3C: National COVID Cohort Collaborative
NAFLD: non-alcoholic fatty liver disease
NCATS: National Center for Advancing Translational Sciences
NIH: National Institutes of Health
NREVSS: National Respiratory and Enteric Virus Surveillance System
OHDSI: Observational Health Data Sciences and Informatics Network
OMOP: Observational Medical Outcomes Partnership
SARS-CoV-2: Severe Acute Respiratory Syndrome Coronavirus 2
SNOMED: Standard Nomenclature of Medicine

## Acknowledgements

The analyses described in this publication were conducted with data or tools accessed through the NCATS N3C Data Enclave covid.cd2h.org/enclave and supported by NCATS U24 TR002306. This research was possible because of the patients whose information is included within the data from participating organizations (covid.cd2h.org/dtas) and the organizations and scientists (covid.cd2h.org/duas) who have contributed to the on-going development of this community resource (cite this https://doi.org/10.1093/jamia/ocaa196).

The N3C data transfer to NCATS is performed under a Johns Hopkins University Reliance Protocol # IRB00249128 or individual site agreements with NIH. The N3C Data Enclave is managed under the authority of the NIH; information can be found at https://ncats.nih.gov/n3c/resources.

We gratefully acknowledge contributions from the following N3C core teams:

- Principal Investigators: Melissa A. Haendel*, Christopher G. Chute*, Kenneth R. Gersing, Anita Walden
- Workstream, subgroup and administrative leaders: Melissa A. Haendel*, Tellen D. Bennett, Christopher G. Chute, David A. Eichmann, Justin Guinney, Warren A. Kibbe, Hongfang Liu, Philip R.O. Payne, Emily R. Pfaff, Peter N. Robinson, Joel H. Saltz, Heidi Spratt, Justin Starren, Christine Suver, Adam B. Wilcox, Andrew E. Williams, Chunlei Wu
- Key liaisons at data partner sites
- Regulatory staff at data partner sites
- Individuals at the sites who are responsible for creating the datasets and submitting data to N3C
- Data Ingest and Harmonization Team: Christopher G. Chute*, Emily R. Pfaff*, Davera Gabriel, Stephanie S. Hong, Kristin Kostka, Harold P. Lehmann, Richard A. Moffitt, Michele Morris, Matvey B. Palchuk, Xiaohan Tanner Zhang, Richard L. Zhu
- Phenotype Team (Individuals who create the scripts that the sites use to submit their data, based on the COVID and Long COVID definitions): Emily R. Pfaff*, Benjamin Amor, Mark M. Bissell, Marshall Clark, Andrew T. Girvin, Stephanie S. Hong, Kristin Kostka, Adam M. Lee, Robert T. Miller, Michele Morris, Matvey B. Palchuk, Kellie M. Walters
- Project Management and Operations Team: Anita Walden*, Yooree Chae, Connor Cook, Alexandra Dest, Racquel R. Dietz, Thomas Dillon, Patricia A. Francis, Rafael Fuentes, Alexis Graves, Andrew J. Neumann, Shawn T. O’Neil, Andréa M. Volz, Elizabeth Zampino
- Partners from NIH and other federal agencies: Christopher P. Austin*, Kenneth R. Gersing*, Samuel Bozzette, Mariam Deacy, Nicole Garbarini, Michael G. Kurilla, Sam G. Michael, Joni L. Rutter, Meredith Temple-O’Connor
- Analytics Team (Individuals who build the Enclave infrastructure, help create codesets, variables, and help Domain Teams and project teams with their datasets): Benjamin Amor*, Mark M. Bissell, Katie Rebecca Bradwell, Andrew T. Girvin, Amin Manna, Nabeel Qureshi

The authors thank the Publication Committee for their review of this publication to ensure compliance with ICMJE guidelines, the N3C User Code of Conduct, and appropriate author attribution.

- Publication Committee Review Team: Carolyn Bramante, Jeremy R. Harper, Wendy Hernandez, Farrukh M. Koraishy, Saidulu Mattapally, Amit Saha, Satyanarayana Vedula

Stony Brook University — U24TR002306, University of Oklahoma Health Sciences Center — U54GM104938: Oklahoma Clinical and Translational Science Institute (OCTSI), West Virginia University — U54GM104942: West Virginia Clinical and Translational Science Institute (WVCTSI), University of Mississippi Medical Center — U54GM115428: Mississippi Center for Clinical and Translational Research (CCTR), University of Nebraska Medical Center — U54GM115458: Great Plains IDeA-Clinical & Translational Research, Maine Medical Center — U54GM115516: Northern New England Clinical & Translational Research (NNE-CTR) Network, Wake Forest University Health Sciences — UL1TR001420: Wake Forest Clinical and Translational Science Institute, Northwestern University at Chicago — UL1TR001422: Northwestern University Clinical and Translational Science Institute (NUCATS), University of Cincinnati — UL1TR001425: Center for Clinical and Translational Science and Training, The University of Texas Medical Branch at Galveston — UL1TR001439: The Institute for Translational Sciences, Medical University of South Carolina — UL1TR001450: South Carolina Clinical & Translational Research Institute (SCTR), University of Massachusetts Medical School Worcester — UL1TR001453: The UMass Center for Clinical and Translational Science (UMCCTS), University of Southern California — UL1TR001855: The Southern California Clinical and Translational Science Institute (SC CTSI), Columbia University Irving Medical Center — UL1TR001873: Irving Institute for Clinical and Translational Research, George Washington Children’s Research Institute — UL1TR001876: Clinical and Translational Science Institute at Children’s National (CTSA-CN), University of Kentucky — UL1TR001998: UK Center for Clinical and Translational Science, University of Rochester — UL1TR002001: UR Clinical & Translational Science Institute, University of Illinois at Chicago — UL1TR002003: UIC Center for Clinical and Translational Science, Penn State Health Milton S. Hershey Medical Center — UL1TR002014: Penn State Clinical and Translational Science Institute, The University of Michigan at Ann Arbor — UL1TR002240: Michigan Institute for Clinical and Health Research, Vanderbilt University Medical Center — UL1TR002243: Vanderbilt Institute for Clinical and Translational Research, University of Washington — UL1TR002319: Institute of Translational Health Sciences, Washington University in St. Louis — UL1TR002345: Institute of Clinical and Translational Sciences, Oregon Health & Science University — UL1TR002369: Oregon Clinical and Translational Research Institute, University of Wisconsin-Madison — UL1TR002373: UW Institute for Clinical and Translational Research, Rush University Medical Center — UL1TR002389: The Institute for Translational Medicine (ITM), The University of Chicago — UL1TR002389: The Institute for Translational Medicine (ITM), University of North Carolina at Chapel Hill — UL1TR002489: North Carolina Translational and Clinical Science Institute, University of Minnesota — UL1TR002494: Clinical and Translational Science Institute, Children’s Hospital Colorado — UL1TR002535: Colorado Clinical and Translational Sciences Institute, The University of Iowa — UL1TR002537: Institute for Clinical and Translational Science, The University of Utah — UL1TR002538: Uhealth Center for Clinical and Translational Science, Tufts Medical Center — UL1TR002544: Tufts Clinical and Translational Science Institute, Duke University — UL1TR002553: Duke Clinical and Translational Science Institute, Virginia Commonwealth University — UL1TR002649: C. Kenneth and Dianne Wright Center for Clinical and Translational Research, The Ohio State University — UL1TR002733: Center for Clinical and Translational Science, The University of Miami Leonard M. Miller School of Medicine — UL1TR002736: University of Miami Clinical and Translational Science Institute, University of Virginia — UL1TR003015: iTHRIVL Integrated Translational health Research Institute of Virginia, Carilion Clinic — UL1TR003015: iTHRIVL Integrated Translational health Research Institute of Virginia, University of Alabama at Birmingham — UL1TR003096: Center for Clinical and Translational Science, Johns Hopkins University — UL1TR003098: Johns Hopkins Institute for Clinical and Translational Research, University of Arkansas for Medical Sciences — UL1TR003107: UAMS Translational Research Institute, Nemours — U54GM104941: Delaware CTR ACCEL Program, University Medical Center New Orleans — U54GM104940: Louisiana Clinical and Translational Science (LA CaTS) Center, University of Colorado Denver, Anschutz Medical Campus — UL1TR002535: Colorado Clinical and Translational Sciences Institute, Mayo Clinic Rochester — UL1TR002377: Mayo Clinic Center for Clinical and Translational Science (CCaTS), Tulane University — UL1TR003096: Center for Clinical and Translational Science, Loyola University Medical Center — UL1TR002389: The Institute for Translational Medicine (ITM), Advocate Health Care Network — UL1TR002389: The Institute for Translational Medicine (ITM), OCHIN — INV-018455: Bill and Melinda Gates Foundation grant to Sage Bionetworks, The Rockefeller University — UL1TR001866: Center for Clinical and Translational Science, The Scripps Research Institute — UL1TR002550: Scripps Research Translational Institute, University of Texas Health Science Center at San Antonio — UL1TR002645: Institute for Integration of Medicine and Science, The University of Texas Health Science Center at Houston — UL1TR003167: Center for Clinical and Translational Sciences (CCTS), NorthShore University HealthSystem — UL1TR002389: The Institute for Translational Medicine (ITM), Yale New Haven Hospital — UL1TR001863: Yale Center for Clinical Investigation, Emory University — UL1TR002378: Georgia Clinical and Translational Science Alliance, Weill Medical College of Cornell University — UL1TR002384: Weill Cornell Medicine Clinical and Translational Science Center, Montefiore Medical Center — UL1TR002556: Institute for Clinical and Translational Research at Einstein and Montefiore, Medical College of Wisconsin — UL1TR001436: Clinical and Translational Science Institute of Southeast Wisconsin, University of New Mexico Health Sciences Center — UL1TR001449: University of New Mexico Clinical and Translational Science Center, George Washington University — UL1TR001876: Clinical and Translational Science Institute at Children’s National (CTSA-CN), Stanford University — UL1TR003142: Spectrum: The Stanford Center for Clinical and Translational Research and Education, Regenstrief Institute — UL1TR002529: Indiana Clinical and Translational Science Institute, Cincinnati Children’s Hospital Medical Center — UL1TR001425: Center for Clinical and Translational Science and Training, Boston University Medical Campus — UL1TR001430: Boston University Clinical and Translational Science Institute, The State University of New York at Buffalo — UL1TR001412: Clinical and Translational Science Institute, Aurora Health Care — UL1TR002373: Wisconsin Network For Health Research, Brown University — U54GM115677: Advance Clinical Translational Research (Advance-CTR), Rutgers, The State University of New Jersey — UL1TR003017: New Jersey Alliance for Clinical and Translational Science, Loyola University Chicago — UL1TR002389: The Institute for Translational Medicine (ITM), #N/A — UL1TR001445: Langone Health’s Clinical and Translational Science Institute, Children’s Hospital of Philadelphia — UL1TR001878: Institute for Translational Medicine and Therapeutics, University of Kansas Medical Center — UL1TR002366: Frontiers: University of Kansas Clinical and Translational Science Institute, Massachusetts General Brigham — UL1TR002541: Harvard Catalyst, Icahn School of Medicine at Mount Sinai — UL1TR001433: ConduITS Institute for Translational Sciences, Ochsner Medical Center — U54GM104940: Louisiana Clinical and Translational Science (LA CaTS) Center, HonorHealth — None (Voluntary), University of California, Irvine — UL1TR001414: The UC Irvine Institute for Clinical and Translational Science (ICTS), University of California, San Diego — UL1TR001442: Altman Clinical and Translational Research Institute, University of California, Davis — UL1TR001860: UCDavis Health Clinical and Translational Science Center, University of California, San Francisco — UL1TR001872: UCSF Clinical and Translational Science Institute, University of California, Los Angeles — UL1TR001881: UCLA Clinical Translational Science Institute, University of Vermont — U54GM115516: Northern New England Clinical & Translational Research (NNE-CTR) Network, Arkansas Children’s Hospital — UL1TR003107: UAMS Translational Research Institute

## Author Contributions

Authorship was determined using ICMJE recommendations.

*Ge*: Study concept and design; data extraction; analysis and interpretation of data; drafting of manuscript; critical revision of the manuscript for important intellectual content; statistical analysis

*Pletcher*: Acquisition of data; interpretation of data; critical revision of the manuscript for important intellectual content

*Lai*: Study concept and design; analysis and interpretation of data; drafting of manuscript; critical revision of the manuscript for important intellectual content; obtained funding; study supervision

## N3C Consortium Collaborators/Authors Contributions

*Harper*: Data quality assurance; consortial governance; N3C phenotype definition

*Chute*: Clinical data model expertise; data curation; data integration; data quality assurance; data security; funding acquisition; governance; critical revision of the manuscript for important intellectual content; N3C Phenotype definition; project management; regulatory oversight / admin

*Haendel*: Funding acquisition; governance; project management; regulatory oversight / admin

## Writing Assistance

None.

