## Supplemental Tables for "Outcomes of SARS-CoV-2 Infection in Patients with Chronic Liver Disease and Cirrhosis: a N3C Study"

**Supplemental Table 1 – Standard OMOP concept identifiers for SARS-CoV-2 testing per N3C shared logic sets**

| SARS-CoV-2 Test Type | OMOP Concept Identifier(s) |
| --- | --- |
| Antibody | 586515, 586521, 586522, 706176, 706177, 706178, 706179, 706181, 723459, 757679, 757680 |
| Culture | 586516 |
| Nucleic Acid Amplification | 586517, 586518, 586519, 586520, 586523, 586526, 706154, 706155, 706156, 706157, 706158, 706159, 706160, 706161, 706163, 706165, 706166, 706167, 706168, 706169, 706170, 706171, 706172, 706173, 706174, 706175, 715260, 715261, 715262, 757677, 757678 |
| Abbreviations: OMOP, Observational Medical Outcomes Partnership. | |

**Supplemental Table 2 – Standard OMOP concept identifiers for chronic liver disease etiologies, alcohol use and its complications, and cirrhosis and its complications**

|  |  | Validated ICD-10-CM Code(s) | OMOP Concept Identifier(s) |
| --- | --- | --- | --- |
| Etiology of Chronic Liver Disease | |  |  |
|  | Non-Alcoholic Fatty Liver Disease^19,20,26^ | K76.0 without an associated alcohol use ICD10*, K75.81 | 4059290 without an associated alcohol use concept ID*, 40484532 |
|  | Chronic Hepatitis C^19,20^ | B17.1, B18.2, B19.2 | 192242, 198964, 197494 |
|  | Alcohol-Associated Liver Disease^19,20,24–26,36^ | K70.0, K70.1, K70.2, K70.3, K70.4, K70.9, and K76.0 with an associated alcohol use ICD-10 code* | 4340383, 4340385, 196463, 4340386, 201612, and 4059290 with an associated alcohol use concept ID* |
|  | Chronic Hepatitis B^19,20^ | B16.X, B17,0, B18.0, B18.1, B19.1 | 197795, 197493, 192240, 439674, 4281232 |
|  | Cholestatic Liver Disease^21^ | K74.3, K74.4, K74.5, K83.01 | 4135822, 4046123, 192675, 4058821 |
|  | Autoimmune Hepatitis^22^ | K73.2, K75.4 | 4026125, 200762 |
| *Alcohol Use and Its Complications^24–26,36^ | | F10.X, G62.1, G31.2, G72.1, I42.6, K29.2, K85.2, K86.0 | 376383, 378421, 36714559, 4078688, 318773, 195300, 4340493, 4340964, 432456, 283761, 37814 |
| Cirrhosis and Its Complications^11,24,27^ | |  | |
|  | Cirrhosis | K70.30, K74.69, K74.60, E83.11, K71.7, K72.1, K74.3, K74.4, K74.5 | 196463, 4064161, 4163735, 4026136, 4340390, 4135822, 4046123, 192675 |
|  | Varices, not bleeding | I85.00, I86.4, I85.1 | 22340, 24966, 4237824, 4111998 |
|  | Varices, bleeding | I85.01, I86.41, I85.11 | 28779, 4087310, 4112183, |
|  | Ascites | K70.31, K70.11, K71.51, R18.8 | 46269816, 46269835, 46273476, 200528 |
|  | Spontaneous Bacterial Peritonitis | K65.2 | 199863 |
|  | Hepatic Encephalopathy | K72.91, G93.40, K72.11, K70.41, K71.11, K72.01, B19.0, B19.11, B19.21 | 4245975, 377604, 372887, 46269836, 46269818, 377604, 196029, 200031, 439672 |
|  | Hepatorenal Syndrome | K76.7 | 196455 |
|  | Hepatopulmonary Syndrome | K76.81 | 4159144 |
| Abbreviations: ICD-10-CM, International Classification of Diseases, Tenth Revision, Clinical Modification; OMOP, Observational Medical Outcomes Partnership. | | | |

**Supplemental Table 3 – Standard OMOP concept identifiers for international normalized ratio (INR)**

| OMOP Concept Identifier(s) | Concept Name(s) |
| --- | --- |
| 3039326 | INR in Platelet poor plasma by Coagulation assay --post heparin neutralization |
| 3022217 | INR in Platelet poor plasma by Coagulation assay |
| 3051593 | INR in Capillary blood by Coagulation assay |
| 3032080 | INR in Blood by Coagulation assay |
| 3042605 | INR in Platelet poor plasma or blood by Coagulation assay |
| 4261078 | Calculation of international normalized ratio |
| Abbreviations: INR, international normalized ratio; OMOP, Observational Medical Outcomes Partnership. | |

**Supplemental Table 4 – Sensitivity Analyses of Cox Regressions Involving Patients with Cirrhosis**

| Model Evaluated | | Number | aHR | 95% CI | P-Value |
| --- | --- | --- | --- | --- | --- |
| SARS-CoV-2 Infection Among Patients with Cirrhosis (Cirrhosis/Positive versus Cirrhosis/Negative) | | | | | |
|  | Multivariate (MV) | 62,191 | 2.43 | 2.23-2.64 | <0.01 |
|  | MV + MELD-Na | 10,816 | 2.02 | 1.75-2.34 | <0.01 |
|  | MV + Serum Albumin | 27,819 | 1.81 | 1.51-1.88 | <0.01 |
|  | MV + MELD-Na + Serum Albumin | 9,774 | 1.81 | 1.56-2.10 | <0.01 |
| Age in Factors Associated with Death Among Cirrhosis/Positive Patients | | | | | |
|  | Multivariate (MV) | 8,135 | 1.03 | 1.03-1.04 | <0.01 |
|  | MV + MELD-Na | 1,307 | 1.04 | 1.03-1.05 | <0.01 |
|  | MV + Serum Albumin | 3,635 | 1.02 | 1.02-1.03 | <0.01 |
|  | MV + MELD-Na + Serum Albumin | 1,200 | 1.04 | 1.03-1.05 | <0.01 |
| Hispanic Ethnicity (Reference White) in Factors Associated with Death Among Cirrhosis/Positive Patients | | | | | |
|  | Multivariate (MV) | 8,135 | 1.24 | 1.00-1.53 | 0.05 |
|  | MV + MELD-Na | 1,307 | 1.11 | 0.80-1.56 | 0.54 |
|  | MV + Serum Albumin | 3,635 | 1.10 | 0.86-1.41 | 0.46 |
|  | MV + MELD-Na + Serum Albumin | 1,200 | 0.97 | 0.68-1.38 | 0.87 |
| Unknown/Other Race (Reference White) in Factors Associated with Death Among Cirrhosis/Positive Patients | | | | | |
|  | Multivariate (MV) | 8,135 | 1.65 | 1.27-2.15 | <0.01 |
|  | MV + MELD-Na | 1,307 | 1.18 | 0.77-1.82 | 0.44 |
|  | MV + Serum Albumin | 3,635 | 1.61 | 1.17-2.20 | <0.01 |
|  | MV + MELD-Na + Serum Albumin | 1,200 | 1.31 | 0.85-2.04 | 0.22 |
| Cholestatic Liver Disease (Reference NAFLD) in Factors Associated with Death Among Cirrhosis/Positive Patients | | | | | |
|  | Multivariate (MV) | 8,135 | 0.63 | 0.42-0.94 | 0.02 |
|  | MV + MELD-Na | 1,307 | 0.37 | 0.17-0.80 | 0.01 |
|  | MV + Serum Albumin | 3,635 | 0.67 | 0.41-1.08 | 0.10 |
|  | MV + MELD-Na + Serum Albumin | 1,200 | 0.54 | 0.25-1.17 | 0.12 |
| Charlson Comorbidity Index as Etiology in Factors Associated with Death Among Cirrhosis/Positive Patients | | | | | |
|  | Multivariate (MV) | 8,135 | 1.04 | 1.02-1.06 | <0.01 |
|  | MV + MELD-Na | 1,307 | 0.99 | 0.96-1.02 | 0.57 |
|  | MV + Serum Albumin | 3,635 | 1.02 | 1.00-1.04 | 0.06 |
|  | MV + MELD-Na + Serum Albumin | 1,200 | 1.01 | 0.98-1.05 | 0.50 |
| Midwest Location (Reference Northeast) as Etiology in Factors Associated with Death Among Cirrhosis/Positive Patients | | | | | |
|  | Multivariate (MV) | 8,135 | 0.60 | 0.45-0.80 | 0.01 |
|  | MV + MELD-Na | 1,307 | 0.74 | 0.47-1.17 | 0.20 |
|  | MV + Serum Albumin | 3,635 | 0.93 | 0.66-1.31 | 0.67 |
|  | MV + MELD-Na + Serum Albumin | 1,200 | 1.08 | 0.65-1.80 | 0.78 |
| Other Location (Reference Northeast) as Etiology in Factors Associated with Death Among Cirrhosis/Positive Patients | | | | | |
|  | Multivariate (MV) | 8,135 | 0.58 | 0.46-0.74 | 0.01 |
|  | MV + MELD-Na | 1,307 | 0.84 | 0.57-1.25 | 0.39 |
|  | MV + Serum Albumin | 3,635 | 0.90 | 0.67-1.20 | 0.46 |
|  | MV + MELD-Na + Serum Albumin | 1,200 | 1.04 | 0.68-1.61 | 0.85 |
| Abbreviations: aHR, adjusted hazard ratio; CI, confidence interval; HR, hazard ratio; MELD-Na, Model for End-Stage Liver Disease-Sodium; MV, multivariate; NAFLD, non-alcoholic fatty liver disease. | | | | | |
